# Primary series and booster COVID-19 vaccine effectiveness in a cohort of healthcare workers in Albania during a BA.1 and BA.2 variant period, January – May 2022

**DOI:** 10.1101/2023.05.05.23289503

**Authors:** Iris Finci, Madelyn Yiseth Rojas Castro, Iris Hasibra, Jonilda Sulo, Albana Fico, Rovena Daja, Adela Vasili, Majlinda Kota, Iria Preza, Barbara Mühlemann, Christian Drosten, Richard Pebody, Kathryn E. Lafond, Esther Kissling, Mark A. Katz, Silvia Bino

## Abstract

**Background:** Healthcare workers (HCWs) have experienced high rates of COVID-19 morbidity and mortality. We estimated COVID-19 two-dose primary series and monovalent booster vaccine effectiveness (VE) against symptomatic SARS-CoV-2 Omicron (BA.1 and BA.2) infection among HCWs in three Albanian hospitals during January–May 2022.

**Methods:** Study participants completed weekly symptom questionnaires, underwent PCR testing when symptomatic, and provided quarterly blood samples for serology. We estimated VE using Cox regression models (1-hazard ratio), with vaccination status as the time-varying exposure and unvaccinated HCWs as the reference group, adjusting for potential confounders: age, sex, prior SARS-CoV-2 infection (detected by PCR, rapid-antigen test or serology), and household size.

**Results:** At the start of the analysis period, 76% of 1,462 HCWs had received a primary series, 10% had received a booster dose, and 9% were unvaccinated; 1,307 (89%) HCWs had evidence of prior infection. Overall, 86% of primary series and 98% of booster doses received were BNT162b2. The median time interval from the second dose and the booster dose to the start of the analysis period was 289 days (IQR:210– 292) and 30 days (IQR:22–46), respectively. VE against symptomatic PCR-confirmed infection was 34% (95%CI: -36;68) for the primary series and 88% (95%CI: 38;98) for the booster.

**Conclusions:** Among Albanian HCWs, most of whom had been previously infected, COVID-19 booster dose offered improved VE during a period of Omicron BA.1 and BA.2 circulation. Our findings support promoting booster dose uptake among Albanian HCWs, which, as of January 2023, was only 20%.

## Introduction

Healthcare workers (HCWs) have suffered considerable morbidity and mortality during the COVID-19 pandemic^1,2^. Vaccinating HCWs against COVID-19 decreases COVID-19-related illness and absenteeism and therefore helps to maintain a functioning health care workforce during periods of high burden on health systems. Additionally, vaccinating HCWs can potentially reduce onward SARS-CoV-2 transmission^3^. Understanding COVID-19 vaccine effectiveness (VE) among HCWs is critical to inform optimal vaccination policies.

As of February 2023, in the European Region of the World Health Organization (WHO), primary series and booster COVID-19 vaccine uptake among HCWs was considerably lower in the 5/19 middle-income countries (MICs) that reported data to WHO (66% and 27%, respectively) compared to the 20/34 high income countries (HICs) that reported (82% and 59%, respectively)^4^.

Few COVID-19 VE studies have been reported in MICs in Europe or globally, particularly during periods of Omicron circulation^5^. Differences in population health and demographics, differences in health systems and time of access to COVID-19 vaccines between HICs and MICs may variably impact vaccine performance, underscoring the need for VE studies in MICs to guide policy^6^. Furthermore, local data demonstrating positive VE estimates can be helpful in promoting vaccine uptake in countries where vaccine acceptance is low.

In early 2022, the spread of SARS-CoV-2 Omicron, a variant with higher transmissibility compared to previous variants^7^, led to elevated pressure on healthcare systems across Europe, and many infected HCWs missed work^8^. To date, studies of COVID-19 VE against Omicron, mostly from HICs^9^, have shown sustained moderate to high primary series VE against severe disease (76% within 6 months after the last dose) but much lower VE against milder symptomatic infection (35% within 6 months after last dose)^10^; however, VE estimates against mild infection increased to 62%^11^ and 71%^10^ following booster doses.

In Albania, an upper-MIC of 2.8 million inhabitants in Southeast Europe, COVID-19 vaccination with BNT162b2 (Comirnaty, Pfizer-BioNTech) started on 11 January 2021, and HCWs were a high-priority group for vaccination. Additionally, ChAdOx1-S (Vaxzervria, AstraZeneca) and CoronaVac (Sinovac Life Sciences) were introduced in mid-March 2021. On 15 October 2021, the Albanian National Technical Advisory Group for Immunizations recommended a booster vaccine for all HCWs who had completed their primary series at least six months prior. As of 31 January 2023, while most Albanian HCWs (83%) had received primary series vaccine, only 20% had received a booster (third) dose^4^.

In February 2021, we established a cohort of HCWs in Albania to prospectively monitor COVID-19 VE against SARS-CoV-2 infection^12–14^. In this analysis, we evaluated primary series and booster dose VE against SARS-CoV-2 infection during BA.1 and BA.2 Omicron period.

## Methods

### Study Design

We conducted an interim analysis of an ongoing prospective cohort study that started in February 2021. Our objective was to estimate VE of COVID-19 primary vaccine series and booster dose against symptomatic and asymptomatic SARS-CoV-2 infection among HCWs at three hospitals in Albania, from January through May 2022. Study methods and early primary series VE results have been previously published^12,14^.

### Data Collection

At enrolment (February–May 2021), participants completed a questionnaire that included questions about demographics, health status, hospital role and prior COVID-19 vaccination. During the study, participants completed weekly symptom questionnaires; participants who reported having any symptom included in the Albanian MOH COVID-19 case definition^12^ provided a nasopharyngeal specimen that was tested for SARS-CoV-2 by RT-PCR at the Institute of Public Health Laboratory in Tirana, Albania.

Study staff cross-checked the Albanian National Surveillance of Infectious Diseases Electronic Information System (SISI) to identify any PCR or rapid antigen tests (RATs) performed in laboratories and hospitals outside of the study, and entered these data into the study database. Self-tested RAT results were not recorded. Participants who tested positive for SARS-CoV-2 by PCR or RAT were administered a follow-up questionnaire about their course of illness 30 days after their positive test. Study staff verified participants’ COVID-19 vaccination status through the national integrated immunization information system (IIS) and the family care physicians web-based medical data system (E-vizita). All study data were entered securely and stored in REDCap^15^.

### Laboratory procedures

We collected blood specimens from participants at enrolment and then every three months throughout the study. Specimens were tested for total anti-nucleocapsid antibody using the Platelia SARS-CoV-2 Total Antibody Assay (Bio-Rad Laboratories, Hercules, CA). Cut-off values were determined according to instructions from the package insert

PCR-positive specimens were sent to the Institute of Virology – Charité (Berlin, Germany), where a representative subset of specimens based on dates and location and cycle threshold value below 24 was selected to undergo whole genome sequencing (WGS). We also inspected Albanian molecular surveillance data from the Global Initiative on Sharing All Influenza Data (GISAID)^16^.

### Vaccine effectiveness analysis

We measured primary series and booster dose VE compared to unvaccinated participants. Additionally, we measured the relative VE (rVE) of booster dose, comparing HCWs who received primary series and booster dose vaccination with HCWs who received primary series alone. For both VE and rVE, the primary analytic outcome was PCR-confirmed symptomatic SARS-CoV-2 infection. For the booster dose VE analyses, HCWs started contributing person-time only when they were eligible to receive the booster dose. We also measured VE against two secondary outcomes: an outcome of any PCR-or RAT-confirmed symptomatic SARS-CoV-2 infection,, and a combined outcome of symptomatic and asymptomatic SARS-CoV-2 infection, confirmed by one of three tests: PCR, RAT, and/or seroconversion. Because inactivated vaccines can produce anti-nucleocapsid antibodies, we excluded participants who received CoronaVac from the analysis that included seroconversion as an endpoint. We defined a symptomatic SARS-CoV-2 infection as a participant with symptom onset between 14 days before and 4 days after the collection date of a SARS-CoV-2-positive swab. We defined seroconversion as a positive anti-nucleocapsid antibody test preceded by a negative anti-nucleocapsid antibody test 3 months earlier.

For participants who seroconverted without having a positive PCR test or RAT, we estimated the date of infection using two approaches; for participants who reported symptoms on their weekly questionnaire during the period of seroconversion, the inferred date of infection was the date of symptom onset. For those without reported symptoms, we estimated the date of infection as the mid-point between the last negative serological test and the date three weeks before the subsequent positive serological test^17^.

We defined prior infection as a history of a positive PCR test, RAT or anti-nucleocapsid serology test at the start of the analysis period. For each analysis described above, we also conducted stratified analyses to evaluate VE among participants who did and did not have evidence of previous SARS-CoV-2 infection at any point before the study start. We also assessed the combined protective effect of vaccination and prior SARS-CoV-2 infection using four levels of exposures: (i) primary series and no prior infection (reference group), (ii) primary series and prior infection, (iii) booster vaccination without prior infection and (iv) booster vaccination and prior infection. Finally, we assessed the impact of time since vaccination on VE of primary series vaccination compared to unvaccinated.

### Statistical model

We used Cox proportional hazards models to estimate VE as (1–adjusted hazard ratio)*100; therefore all reported VE estimates are adjusted. Vaccination was a time-varying exposure as vaccination status could have changed over time. Thus, the same participant could contribute person-time to more than one exposure category. Calendar time was used as the underlying time in the Cox regression. We used the Schoenfeld residual test to check the proportional hazard assumption. All analyses were performed using R software^18^.

We calculated unadjusted and adjusted hazard ratio (HR) and estimated VE, including hospital as a fixed effect to account for the multi-site design. We considered the following a priori covariates to be added in the multivariable regression model: previous infection, age, sex, household size, any chronic condition. We included the aforementioned confounders that changed the VE estimate by more than 5%. Person-time contribution started either on January 1, 2022 or from the start of time at risk for those with prior SARS-CoV-2 infection (90 days after positive test or inferred infection date); it ended at the first occurrence of any of the following events: 1) the day of the SARS-CoV-2 infection, 2) the day of the last weekly questionnaire before complete loss to follow-up, 3) the day of withdrawal from the study, or 4) the censor date for the analysis period (31 May 2022). Participants who received a second or third vaccine dose were excluded for 14 days, after which they were considered to be fully immunized with their respective dose and added to the corresponding exposure category. Person-time of participants vaccinated with only one dose was excluded from the analysis. For the secondary analysis that included seroconversion as an outcome, person-time also ended on the day of the last serological result or receipt of an inactivated vaccine. We also performed two sensitivity analyses (Supplemental information).

### Ethical considerations

The study protocol was approved by the WHO Research Ethics Review Committee (ERC) (reference number CERC.0097A) and the Albania IPH ERC (reference number 156). This activity was determined to meet the requirements of public health surveillance as defined in 45 CFR 46.102(l) (2) (CDC reference number 0900f3eb81ce0ede). All participants provided written informed consent. The study is registered with clinicaltrials.gov (Identifier NCT04811391).

## Results

### Descriptive characteristics

During the analysis period, 1 January – 31 May 2022, 1,462 of the initial 1,504 HCWs (97%) were still enrolled in the study and were included in the analysis. At the start of the follow-up, the median age was 44 years (IQR:34–53), and 1,151 (79%) were female. Overall, 691 (47%) were nurses or midwives, 297 (20%) were physicians, 194 (13%) were janitorial staff or food workers, and 280 (19%) had other professions (Table 1). At analysis period start, 1,112 (76%) HCWs had received the primary vaccine series only, 146 (10%) had received a booster dose (third dose), 69 (5%) had received one dose, and 135 (9%) were unvaccinated. Altogether, 959/1,112 (86%) of primary vaccine series were BNT162b2 (Comirnaty, Pfizer-BioNTech), 137 (9%) ChAdOx1-S (Vaxzervria, AstraZeneca) and 11 (1%) CoronaVac (Sinovac Life Sciences). Overall, 143/146 (98%) booster doses were BNT162b2. The median time since receiving the second dose until the start of the analysis period was 289 days (IQR: 210–292 days); for the booster dose, it was 30 days (IQR: 22–45 days) (Figure S2). At analysis start, 1,307 (89%) HCWs had evidence of prior infection, of whom 115 (11%) were infected in the previous 3 months, 836 (57%) were infected 3-12 months earlier, and 356 (24%) were infected more than 12 months prior (Table 1). At the analysis period end, 1070 (73%) participants had received primary series vaccine only, 247 (17%) participants had received a booster dose, 98 (7%) remained unvaccinated and 47 (3%) received only one dose (Table S1). No participants received a second booster dose during the study period. Most booster doses [244/247 (98%)] were BNT162b2, and nearly all participants who received a booster [227/247 (92%)] had received primary series BNT162b2.

**Table 1.**
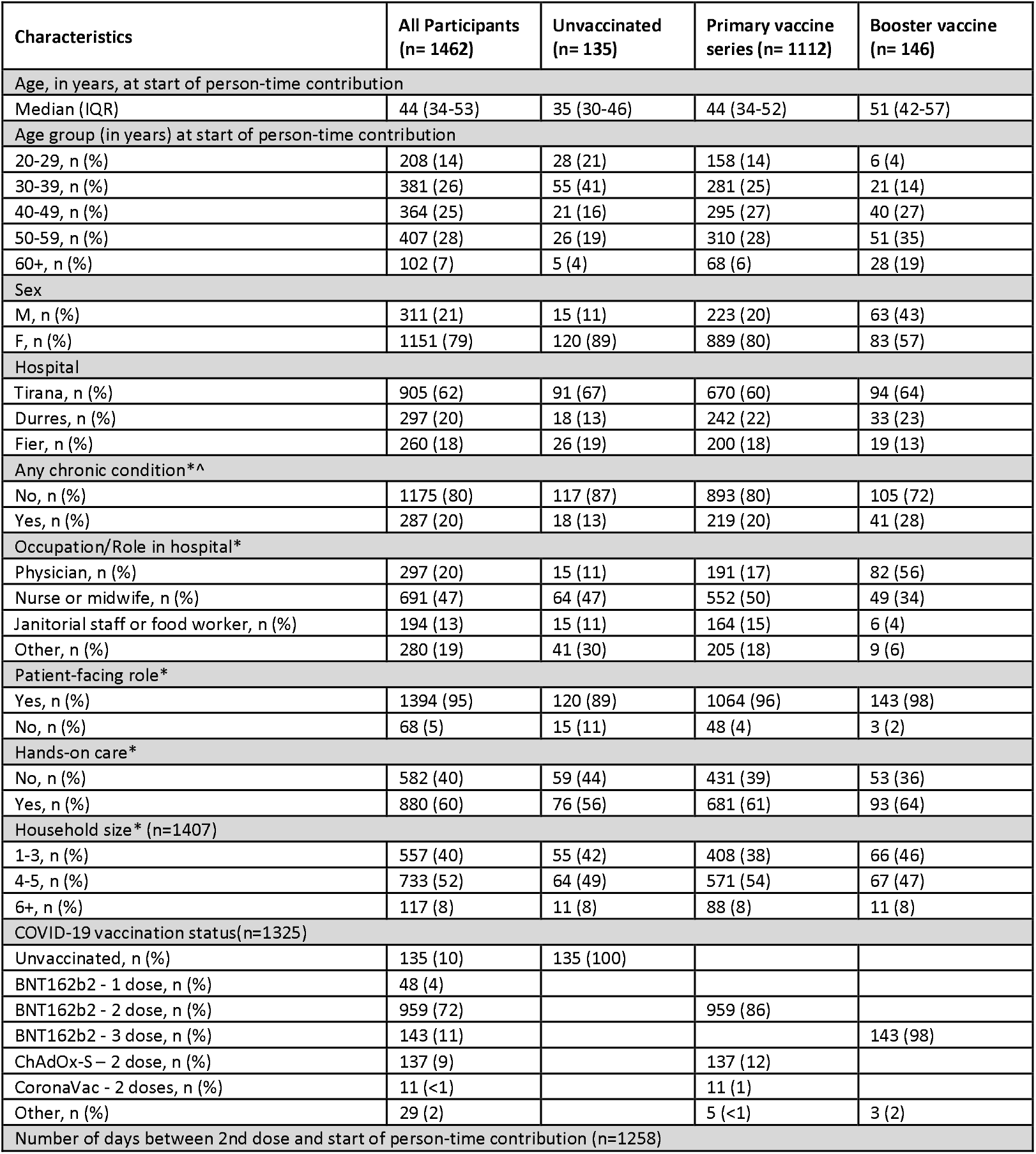
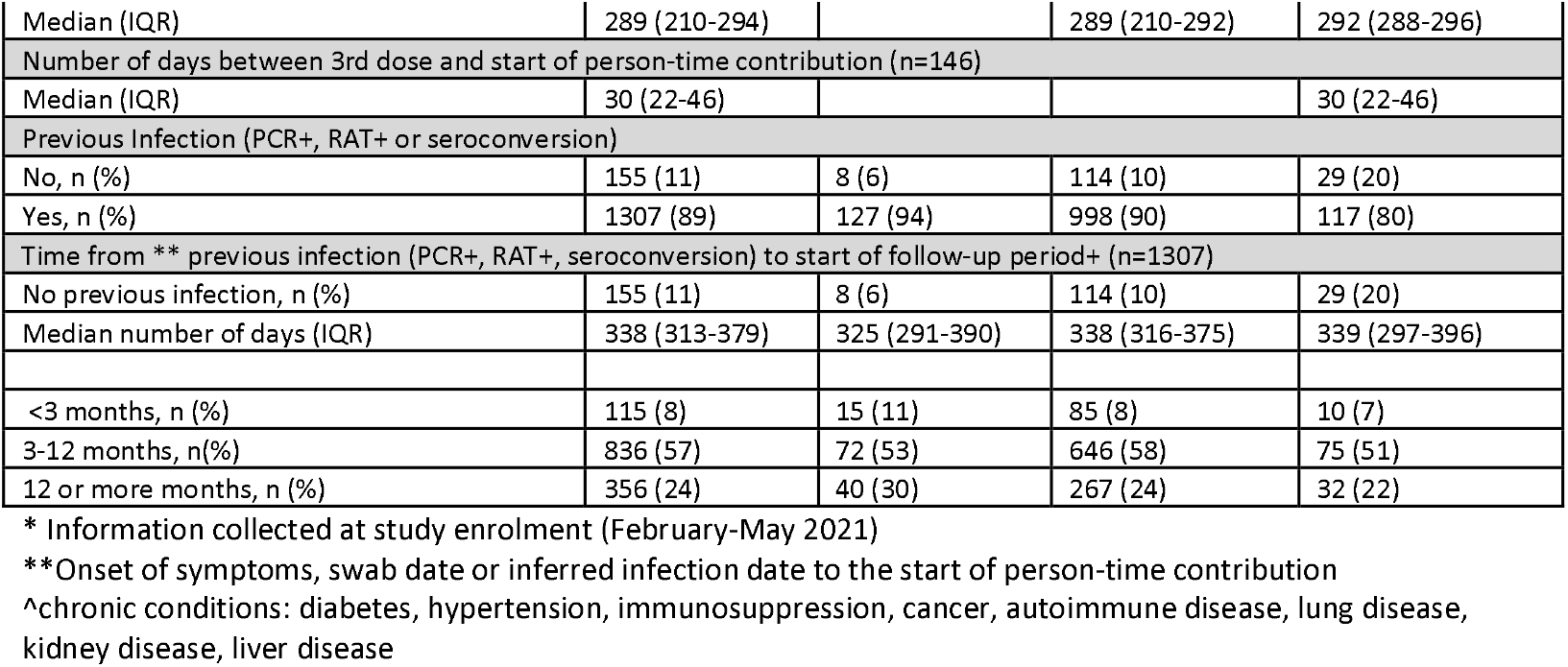
Demographic and occupational characteristics of health care worker participants by vaccination status on the first day of the follow-up period (n=1462), Albania, 2022.

Unvaccinated HCWs were younger (median age: 35 years) and included more females (89%) compared to HCWs that received either a primary vaccine series (median age: 44 years and 80% female) or a booster dose (median age: 51 years and 57% female). Compared to HCWs who received a primary vaccine series only, boosted HCWs were older (median age: 51 years versus 44 years), more often male (43% versus 20%), had a greater proportion with chronic conditions (28% versus 20%), were more often physicians (56% versus 17%) and were less previously infected (80% versus 90%) (Table 1).

### Outcomes

The completion rate of the weekly symptom questionnaire was 89% during the follow-up period. Overall, 167 participants reported symptomatic episodes during the study period; two participants reported two symptomatic episodes, ≥30 days apart. All participants who reported symptoms had a SARS-CoV-2 PCR or RAT test performed; 104 were positive (94 by PCR and 10 by RAT), and 63 participants were negative by PCR or RAT. Among participants included in the analysis, 86 had symptomatic SARS-CoV-2 infections and 107 had asymptomatic infections (55%) (Figure 1B). Of the 86 symptomatic infections, 76 were detected by PCR and 10 by RAT. Of the symptomatic infections, 12 were among unvaccinated participants, 70 were in participants who had received primary vaccine series, and four were in participants who had received a booster dose (Figure 1A). Most of the 107 asymptomatic infections (101 (94%)) were detected through seroconversion (Figure 1B). Combining symptomatic and asymptomatic infections, there were a total of 19 infections among unvaccinated participants, 156 infections in participants with primary vaccine series, and 18 infections in participants who received a booster dose. Almost all symptomatic infections [85/86 (99%)] occurred in January and February, a period when Omicron BA.1 and BA.2 were circulating in Albania (Figure S1B). Overall, 17 (22%) samples underwent genomic sequencing, of which 16 (94%) were of BA.1 or BA.2 Omicron sublineage. BA.1 and BA.2 sublineages accounted for more than 96% of sequenced samples from Albania reported to GISAID in January and February 2022^16^ (Figure S1B).

**Figure 1.**
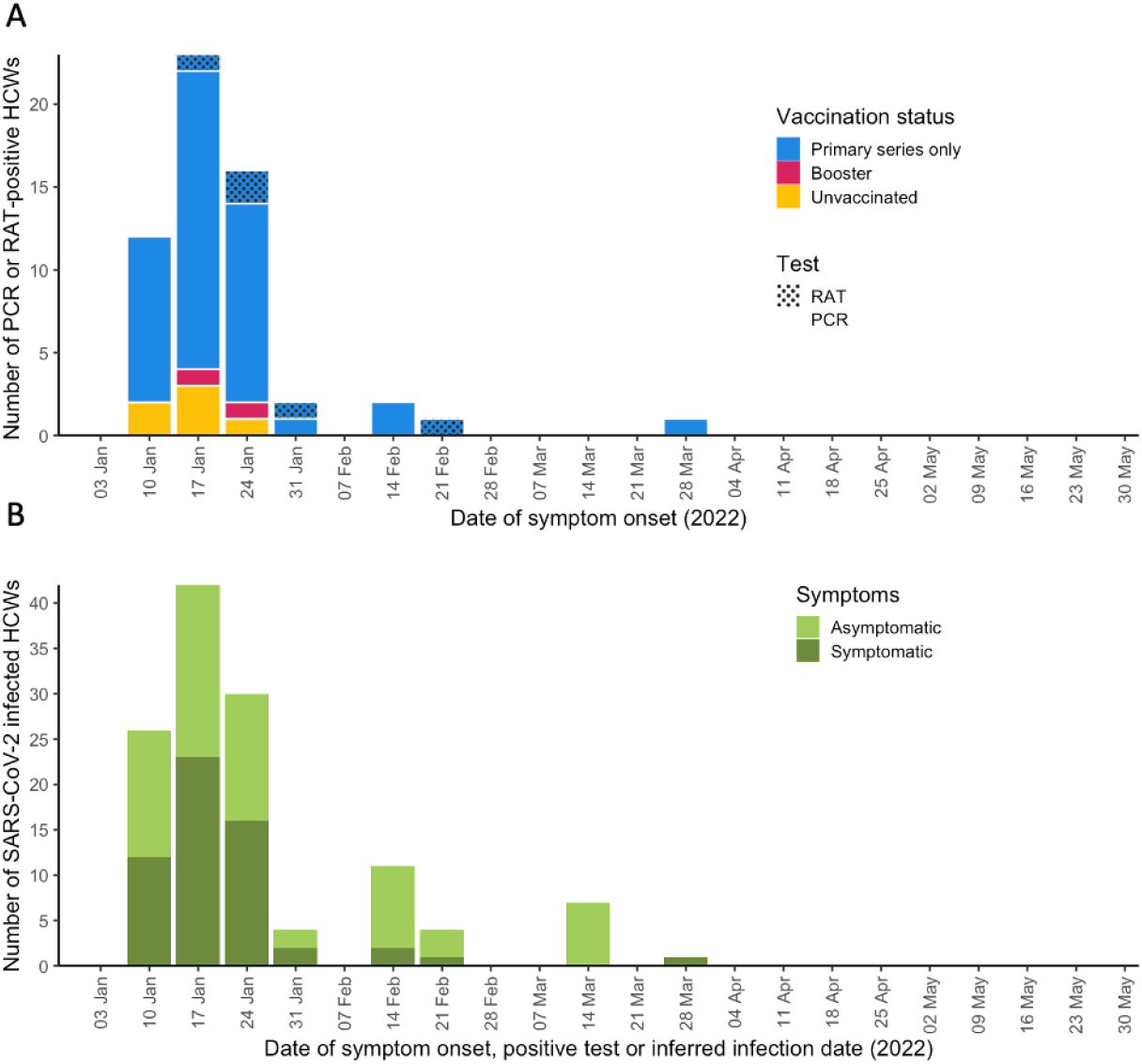
SARS-CoV-2 infections among HCWs by week, January 1 – May 31, 2022. A – Symptomatic SARS-CoV-2 infections stratified by vaccination status and type of confirmatory test, by week; B – all SARS-CoV-2 infections stratified by presence of symptoms, by week

### VE against symptomatic PCR- or RAT-confirmed SARS-CoV-2 infection

VE against symptomatic PCR-confirmed SARS-CoV-2 infection was 34% (95% CI -36;68) for primary vaccine series overall and 26% (95% CI -53;64) for BNT162b2 primary vaccine series (Table 2, Figure 2). Among HCWs with evidence of prior infection, VE of any primary series was 18% (95% CI -83;63) (Table S2). Booster dose VE, compared to unvaccinated HCWs, was 88% (95% CI 39;98) overall and 88% (95% CI 38;98) for participants who had received a BNT162b2 booster. The rVE of a booster dose compared to primary series was 57% (95% CI -25;85) for any booster and 56% (95% CI -28;85) for BNT162b2 boosters compared to BNT162b2 primary series. VE estimates against the combined endpoint of symptomatic PCR- or RAT-confirmed infection were slightly higher with narrower confidence intervals (Figure 2, Table 2). We could not stratify booster VE according to prior infection status due to the small number of events.

**Table 2.**
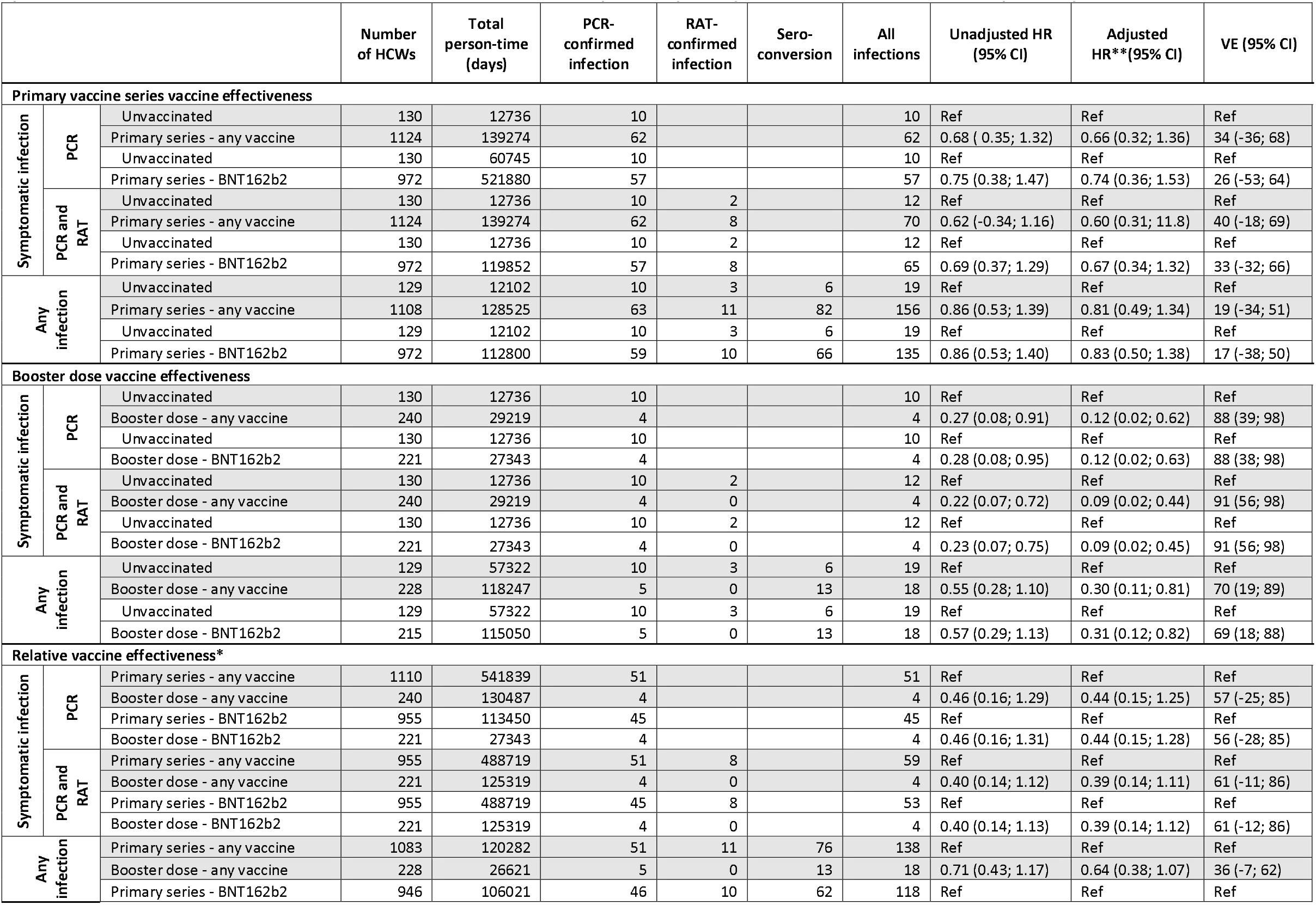
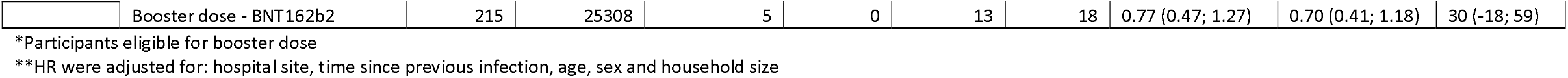
COVID-19 vaccine effectiveness of primary vaccine series and booster dose against SARS-CoV-2 infection during a period of Omicron (BA.1 and BA.2) minance, and relative vaccine effectiveness of booster dose compared to primary vaccine series, Albania, January 1 – May 31, 2022.

**Figure 2.**
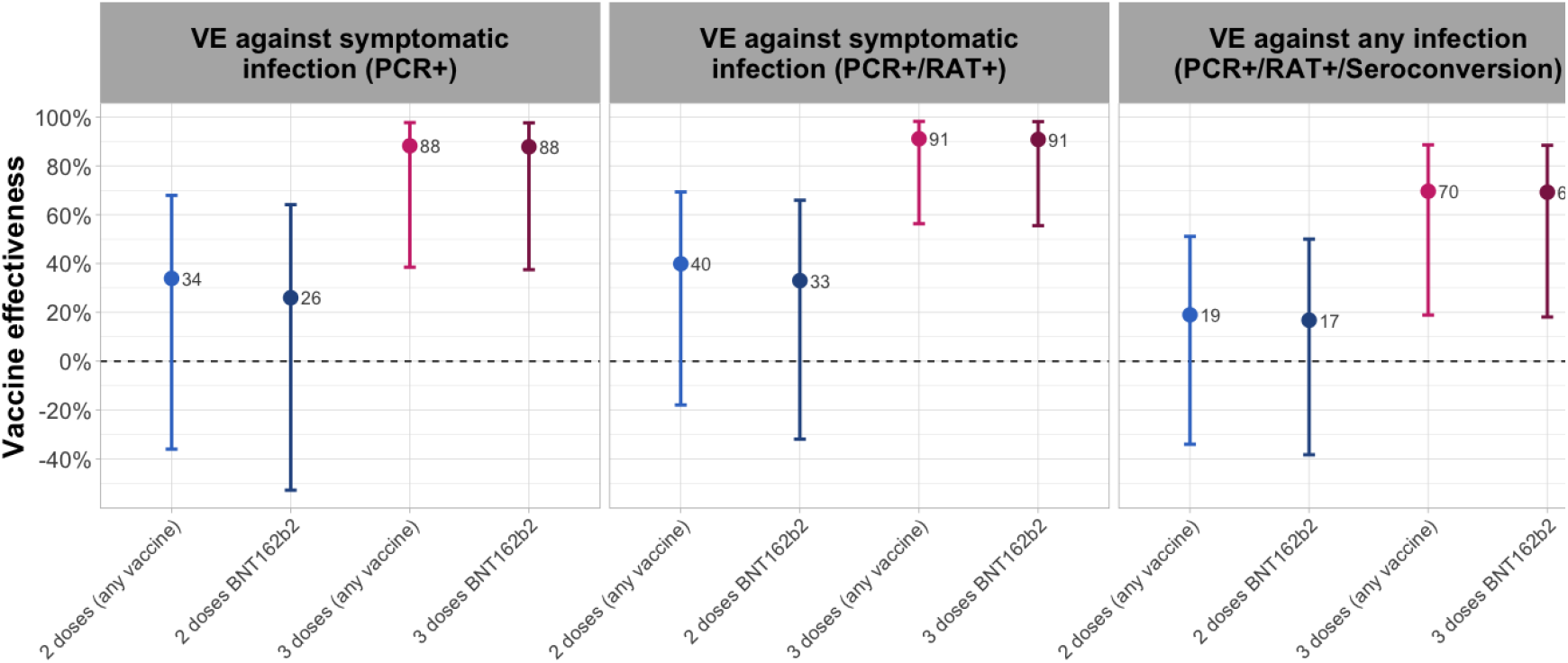
COVID-19 vaccine effectiveness estimates of primary vaccine series and booster dose against PCR-confirmed symptomatic SARS-CoV-2 infection (left), PCR- or RAT-confirmed symptomatic SARS-CoV-2 infection (middle) and any (asymptomatic or symptomatic) SARS-CoV-2 infection confirmed by PCR, RAT or seroconversion (right) Dot – point estimate; lines – 95% confidence interval; blue – Primary series VE; violet – booster dose V

### VE against any (asymptomatic and symptomatic) SARS-CoV-2 infection

Primary series VE against any SARS-CoV-2 infection was 19% (95% CI -34;51) overall and 17% (95% CI 38;50) for BNT162b2 only. Booster dose VE was 70% (95% CI 19;89) overall and 71% (95% CI 18;90) for BNT162b2 only. (Table 2, Figure 2). Among participants eligible for booster vaccination, the rVE of any first booster dose against any SARS-CoV-2 infection was 36% (95% CI -7;62) and the rVE of BNT162b2 booster compared to BNT162b2 primary series was 30% (95% CI -18;59).

Compared to HCWs who received the primary vaccine series and had no evidence of prior infection, rVE of a primary vaccine series combined with prior infection was 79% (95% CI 69;85), rVE of booster dose without a prior infection was 47% (95% CI -21;77) and rVE of a booster dose combined with prior infection was 87% (95% CI 73;94) (Table 3).

**Table 3.**
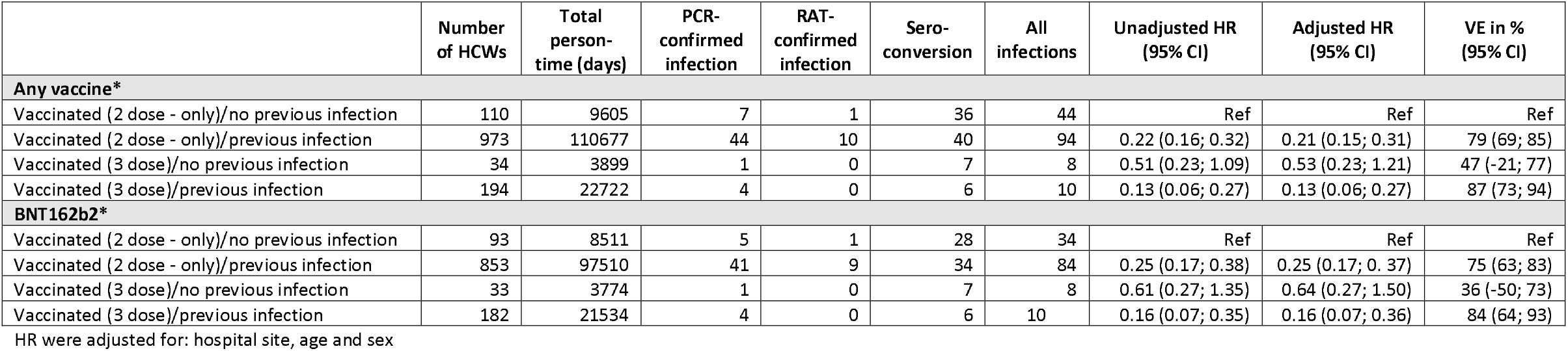
Relative Vaccine effectiveness of booster dose compared to primary vaccine series against any SARS-CoV-2 infection during a period of Omicron and BA.2) predominance, stratified by previous COVID-19 infection, Albania, January 1 – May 31, 2022.

### Change in VE by time since vaccination

For the primary vaccine series, compared to unvaccinated HCWs, VE against symptomatic PCR-confirmed infection was 22% (95% CI -94;69) for participants who had received their second dose within 14-179 days and 41% (95% CI -27;73) for those who had received their second doses ≥180 days. VE against PCR- or RAT-confirmed symptomatic SARS-CoV-2 infection was 33% (95% CI -57;72) within 14-179 days and 43% (95% CI -12;73) ≥180 days (Table 4).

**Table 4.** Vaccine effectiveness against SARS-CoV-2 infection during a period of Omicron (BA.1 and BA.2) predominance by time since receiving primary e series, Albania, January 1 – May 31, 2022.

|  | Number of HCWs | Total person-time (days) | PCR-confirmed infection | RAT-confirmed infection | All infections | Unadjusted HR % (95%CI) | Adjusted HR % (95%CI) | Adjusted VE % (95%CI) |
| --- | --- | --- | --- | --- | --- | --- | --- | --- |
| **Symptomatic SARS-CoV-2-infections detected by PCR by time since vaccination** |  |  |  |  |  |  |  |  |
| Unvaccinated | 130 | 12736 | 10 |  | 10 | Ref | Ref | Ref |
| 2nd dose 14-179 days ago (any vaccine) | 239 | 19124 | 13 |  | 13 | 0.92 (0.39; 2.19) | 0.78 (0.31; 1.94) | 22 (-94; 69) |
| 2nd dose 180+ days ago (any vaccine) | 1018 | 120150 | 49 |  | 49 | 0.64 (0.31; 1.35) | 0.59 (0.27; 1.27) | 42 (-27; 73) |
| **Symptomatic SARS-CoV-2 infection detected by PCR or RAT by time since vaccination** |  |  |  |  |  |  |  |  |
| Unvaccinated | 130 | 12736 | 10 | 2 | 12 | Ref | Ref | Ref |
| 2nd dose 14-179 days ago (any vaccine) | 239 | 14031 | 13 | 0 | 13 | 0.76 (0.33; 1.73) | 0.67 (0.28; 1.57) | 34 (-57; 72) |
| 2nd dose 180+ days ago (any vaccine) | 1018 | 105821 | 49 | 8 | 57 | 0.59 (0.32; 1.12) | 0.57 (0.27; 1.13) | 43 (-13; 73) |

HR were adjusted for: hospital site, previous infection, age and sex

## Discussion

Using data from an ongoing cohort study of HCWs in Albania^12,14^, we found that COVID-19 booster dose VE against symptomatic SARS-CoV-2 infection during a period of Omicron BA.1 and BA.2 predominance was high, at 88%. Conversely, primary vaccine series VE was lower (34%) with low precision. In our cohort, only 17% of HCWs had received a booster dose at the end of the follow-up period, and as of mid-January 2023 only 20% of all HCWs in Albania had received a COVID-19 booster dose^4^. Our high booster VE findings, which to our knowledge reflect the first published booster dose VE results in Southeast Europe, provide evidence to support increased uptake of booster doses among HCWs in Albania.

The high booster dose VE against symptomatic infection, reflecting mainly BNT162b2 booster doses given within one month before the start of follow-up, is slightly higher than findings from studies in the United Kingdom^11^ and Qatar^19^, which found recently administered BNT162b2 booster VE of 67% and 52%, respectively, against symptomatic Omicron BA.1 and BA.2 infection, with confidence intervals that overlap with those from our study. We also found that rVE was 57% for booster dose compared to HCWs who had received the primary vaccine series a median time of 10 months prior to follow-up. The 95% confidence intervals around rVE estimate were wide, with lower bounds that crossed zero. The limited precision of our VE estimates may reflect the relatively small number of events in this analysis. Studies of booster dose rVE against symptomatic Omicron infection in the US^20^ and Qatar^21^ found low to moderate VE, but rVE was much higher against severe outcomes such as hospitalization and death.

In our study, we found that primary vaccine series provided low VE (34%) against symptomatic Omicron infection, although precision was low due to small numbers. This estimate differs from the higher primary series VE estimate we previously found against symptomatic infection during a Delta-predominant period (67%) in the same cohort^14^. In our study, median time since receiving the second dose was almost 10 months; therefore, in addition to the increased immune escape of Omicron, waning of vaccine immunity likely contributed to low primary series VE. Waning VE against symptomatic Omicron infection has been shown in other studies.^9^ In the UK^11^, BNT162b2 VE dropped from 66% in the first month after completion of the primary series to 9% six months after. In our study, we could not observe significant waning primary series immunity, likely due to the low power secondary to the low number of events.

We found that VE was highest among individuals who were both boosted and had prior SARS-CoV-2 infection. These findings are consistent with many published studies that have demonstrated the benefits of hybrid immunity,^22^ and support the Albanian Ministry of Health and Social Protection recommendations that individuals should receive a primary vaccine series regardless of prior infection history, and that HCWs and other priority populations should receive a booster even if they were previously infected. This message is particularly important in the context of populations with high rates of previous infection; a study in the UK reported lower vaccine acceptance among individuals who were previously infected and thought the vaccine would no longer be useful^23^.

This study has several strengths. First, regular serological testing of HCWs allowed us to detect seroconversions, infer asymptomatic infections and estimate VE against any SARS-CoV-2 infection. Serology testing also increased our sensitivity in detecting prior infection. Additional strengths of the study are its prospective cohort design and the very high retention rate of the participants after one year of follow-up. Also, the completion rate of the weekly symptom questionnaire remained high during the follow-up period, and all participants with symptoms were tested for SARS-CoV-2.

Our study has several limitations. Due to the high coverage of the primary vaccine series in our cohort, only a small number of unvaccinated HCWs were available for a comparison group, and this group may have been different from vaccinated participants with respect to their exposures to SARS-CoV-2 and disease risk^25^. To overcome this limitation, we also calculated relative VE of booster dose compared to primary vaccine series. In addition, our study was not powered to estimate VE against severe but rare outcomes like hospitalisation or death. Other studies have shown high, durable primary series VE against severe outcomes during Omicron for BNT162b2^9,10^. Although we were able to detect prior infections through multiple testing platforms, we were not able to analyse time since previous infection. We could not confirm through WGS that all positive participants were infected with BA.1 or BA.2 SARS-CoV-2 Omicron sublineage. There were limited molecular surveillance data from Albania from the first two weeks of January 2022. Finally, because of the low number of events and the relatively low VE, some of our VE estimates had wide confidence intervals and overlapped zero.

In our cohort study of Albanian HCWs we found that a monovalent COVID-19 booster dose offered better protection against symptomatic SARS-CoV-2 infection compared to primary vaccine series during a period of BA.1/BA.2 Omicron circulation. These findings underscore the importance of the Albanian Ministry of Health and Social Protection recommendation that all HCWs should receive a booster dose six months following receipt of their primary vaccine series. Our results, which reflect the first locally generated booster VE data, could be communicated to Albanian HCWs in order to decrease vaccine complacency and increase booster uptake. Our findings could be further generalised and used to promote booster dose uptake among HCWs in neighbouring countries in Southeast Europe and in the general population, where booster uptake remains low.

## Data Availability

All data produced in the present study are available upon reasonable request to the authors.

## Acknowledgments

Jörn Beheim-Schwarzbach, Victor M. Corman, and Terry C. Jones (Institute of Virology, Charité-Universitätsmedizin Berlin, corporate member of Freie Universität Berlin, Humboldt – Universität zu Berlin, and Berlin Institute of Health, Berlin, Germany; and German Centre for Infection Research (DZIF), partner site Charité, Berlin, Germany); Marta Valenciano and Alain Moren (Epiconcept, France).

## Disclaimer

The findings and conclusions in this report are those of the author(s) and do not necessarily represent the views of the Centers for Disease Control and Prevention. The authors affiliated with the World Health Organization (WHO) are alone responsible for the views expressed in this publication and they do not necessarily represent the decisions or policies of the WHO.

## Author contributions

SB, MAK and RP conceived the cohort study on which this analysis is based; IP, IH, JS, AF, RD, AV, MK, MAK, and SB planned and implemented the study, including development of study protocols, data quality checks and acquisition of data; IF, MAK, and SB conceived the article; IF drafted the manuscript and performed the literature search. MYRC performed the data analysis with support from EK. JS, MYRC, and EK directly accessed and verified the raw data and take responsibility for the integrity and accuracy of the analyses. IH, BM, and CD performed lab analysis of study samples. All authors contributed to the interpretation of the results and critically revised the manuscript. All authors had full access to all the data reported in the study and accept responsibility to submit the paper for publication.

## Financial statement

This study was funded by Task Force for Global Health and World Health Organisation Regional Office for Europe.

## Conflict of interest

None of the authors have conflict of interest.

This work has been presented as oral presentation at European Scientific Conference on Applied Infectious Disease Epidemiology in November 2022, Stockholm, Sweden.

